# Diagnosing and staging epithelial ovarian cancer by serum glycoproteomic profiling

**DOI:** 10.1101/2023.03.20.23287422

**Authors:** Chirag Dhar, Prasanna Ramachandran, Gege Xu, Chad Pickering, Tomislav Čaval, Rachel Rice, Bo Zhou, Apoorva Srinivasan, Itati Hundal, Robert Cheng, Paul Aiyetan, Chih-Wei Chu, Thomas J. Herzog, Alexander Babatunde Olawaiye, Gregg Czerwieniec, Francis Jacob, Daniel Serie, Klaus Lindpaintner, Flavio Schwarz

## Abstract

Minimally invasive technologies for early diagnosis of epithelial ovarian cancer (EOC) remain an unmet clinical need. CA-125, a tumor marker secreted into the circulation, is utilized to monitor treatment response and disease relapse in EOC, but has limited utility in accurately triaging patients with pelvic masses of unknown histology. To address this unmet need, we applied a novel blood-based glycoproteomic platform that relies on mass spectrometry coupled to machine learning tools, and identified glycopeptide biomarkers that differentiate between patients with benign pelvic masses and malignant EOC. We then used a subset of these markers to generate a classifier that discriminated between benign pelvic tumors and EOC with sensitivity and specificity of 83.5% and 90.1% in the training set and 86.7 and 86.7% in the testing set, respectively. On subgroup analyses, we noticed that patients with malignant EOC had higher levels of fucosylated markers, primarily of hepatic origin. Furthermore, patients with late-stage EOC (FIGO stage III and IV) had markedly higher levels of tri- and tetra-antennary glycopeptide markers containing fucose. We used these markers to build an independent algorithm that can differentiate between early- and late-stage EOC. Lastly, we detected a similar upregulation of fucosylated glycans and gene expression signatures suggestive of multi-antennary glycans in late-stage EOC tissues. We posit that common mechanisms - possibly driven by cytokines - affect both the tumor glycocalyx and liver-derived glycoproteins. In summary, we generated blood glycoproteomic profiles resemblant of distinct tumor states and identified biomarkers that differentiate between benign and malignant pelvic masses, and/or between early- and late-stage EOC. We also provide mechanistic insights suggesting a direct link between the tumor site and the circulating glycoproteome. These data may inform the development of robust clinical tests to diagnose and stage patients with EOC.

## Introduction

Epithelial ovarian cancer (EOC) is the second-most common gynecologic malignancy, the leading cause of death among gynecological cancers, and the fourth-leading cause of cancer-related death in women in the United States. Like most cancers identified at an early stage, EOC can be treated effectively with surgery and adjuvant therapies (1). However, diagnosis of early-stage disease is challenging due to the nature of the initial clinical signs and symptoms that are subtle and nonspecific, including pelvic pain, urinary urgency and frequency, abdominal bloating, early satiety, loss of appetite, and weight loss. More evident symptoms associated with the development of a space-occupying process generally occur with advanced stage disease (1). Therefore, only 15-20% of EOC cases are diagnosed at an early stage, when the 5-year survival is greater than 90% (2), and the majority are diagnosed at late-stage (FIGO stage III or IV), with 5-year survival rates ranging between 17% and 39% (3).

In addition to severely compromised treatment of serious disease as a consequence of late diagnosis, there is significant over-treatment of benign disease due to lack of sensitive tests to determine the nature of pelvic masses prior to surgical resection: while over 90% of women presenting with a pelvic mass ultimately undergo surgery, only about 20% are found to have malignant disease(4). Moreover, up to 30% of apparent early-stage EOC is upstaged after surgical resection and pathology (5,6). Hence, guidelines issued by the National Comprehensive Cancer Network and the American College of Obstetricians and Gynecologists recommend consideration of diagnostic modalities, including imaging and biochemical tests, to attempt to determine if surgical intervention is indicated.

Circulating biomarkers are used to diagnose ovarian cancer in combination with other tests (7), many of which utilize glycoproteins (8). Elevated levels of CA-125, a glycosylated protein found in blood and on the surface of some ovarian cancer cells (9), can indicate ovarian cancer (10). Its utility is limited by its poor sensitivity and specificity (11). In fact, serum CA-125 is not increased in 21% of ovarian carcinomas; on the other hand, elevated CA-125 levels have been observed in a variety of other malignant and non-malignant conditions such as endometriosis, uterine fibroids, or even during menstruation or pregnancy (12). Therefore, CA-125 is more effective as a tool for monitoring cancer progression and treatment response (13). Tests that monitor CA-125 in combination with other parameters, such as ROCA (14), OvaCheck (15), OVASure (16), ROMA (17), RMI (18), and HE4 (19) have been developed. While these tests generally perform better than CA-125 alone, none of them has found broad acceptance due to their complexity or inadequate performance (16,20–23). The OVA-1 test (24,25), while representing a potential improvement, has not seen significant acceptance by the medical community. Hence, there is still a clear, unfulfilled need for noninvasive diagnostics in EOC with high sensitivity and specificity. Other blood markers such as CA19-9 and CEA have been suggested as potential indicators of distinct types of ovarian cancer (7) but have limited utility.

To address this unmet clinical need, we utilized novel workflows to probe the glycoproteome in serum of ovarian cancer patients. Specifically, we used a platform that combines liquid chromatography-mass spectrometry (LC-MS) glycoproteomic analysis and artificial intelligence algorithms. We identified critical glycopeptide markers that were differentially expressed and built classifiers that differentiated between benign and malignant pelvic masses, as well as early- and late-stage stage EOC. We then detected comparable variations in glycosylation profiles at the tumor site and investigated if common upstream mechanisms led to the observed changes both at the disease site and in circulation. We propose that diagnostic tests based on glycopeptide biomarkers derived from liquid biopsies may be an effective platform for triaging pelvic masses and staging of EOC.

## Methods

### Human subject samples

Serum samples from women with benign tumors (n=151) or with malignant EOC (n=145), and from healthy controls (n=55) were acquired from Indivumed (Hamburg, Germany). Samples were collected prior to therapeutic intervention and in accordance with applicable guidelines and regulations for human subjects’ protection. Information on the FIGO stage of the EOC was available for 98 of the 145 patients (Table 1). Assessment of benign and malignant tumors was based on histopathological analysis of tissue specimens. All samples were stored at -80°C until tested.

**Table 1:** Clinical characteristics of patients included in this study.

|  |  |  | N |  | Median age<br>(in years) |
| --- | --- | --- | --- | --- | --- |
| Healthy controls |  |  | 55 |  | 52 |
| Benign ovarian tumors |  |  | 151 |  | 60 |
| EOC |  |  | 145 |  | 66 |
|  | Early-stage | FIGO Stage I |  | 12 |  |
|  |  | FIGO Stage II |  | 6 |  |
|  | Late-stage | FIGO Stage III |  | 68 |  |
|  |  | FIGO Stage IV |  | 12 |  |
|  | Undocumented |  |  | 47 |  |

For both models an 80/20 train/test split was used.

### Chemicals and reagents

Human serum, dithiothreitol (DTT), and iodoacetamide (IAA) were purchased from MilliporeSigma (St. Louis, MO, USA). Sequencing-grade trypsin was from Promega (Madison, WI, USA), LC-MS-grade water and acetonitrile were from Honeywell (Muskegon, MI, USA), and LC-MS grade formic acid was from Thermo Scientific (Waltham, MA, USA). Stable isotope-labeled peptide internal standards were purchased from Vivitide (Gardner, MA).

### Preanalytical sample preparation

Serum samples were treated with DTT and IAA to reduce and alkylate disulfide bonds, followed by digestion with trypsin at 37°C for 18 hours. The digestion was quenched by adding formic acid to a final concentration of 1% (v/v), followed by addition of a cocktail of stable isotope-labeled peptide internal standards at known concentrations.

### LC-MS analysis

Digested serum samples were separated over an Agilent ZORBAX Eclipse Plus C18 column (2.1 mm × 150 mm i.d., 1.8 μm particle size) using an Agilent 1290 Infinity UHPLC system. The mobile phase A consisted of 3% acetonitrile, 0.1% formic acid in water (v/v), and the mobile phase B of 90% acetonitrile 0.1% formic acid in water (v/v), with the flow rate set at 0.5 mL/minute. After electrospray ionization, operated in positive ion mode, samples were injected into an Agilent 6495B triple quadrupole MS operated in dynamic multiple reaction monitoring (dMRM) mode. Samples were injected in a randomized order with respect to the clinical features and interspersed with reference samples.

### Glycopeptide and peptides quantification

PeakBoundaryNet21, an in-house-developed spectrogram feature recognition and integration software based on recurrent neural networks was used to integrate chromatogram peaks and to obtain molecular abundance quantification for each peptide and glycopeptide (26). R Libraries ‘stats’ and ‘caret’ and python library Scikit-learn (https://scikit-learn.org/stable/) was used for all statistical analyses and for building machine learning models.

A total of 653 peptides and glycopeptides derived from 71 high-abundance (concentrations of ≥10 μg/ml) serum glycoproteins were quantified. Intensity of glycopeptides and peptides were corrected for within-run drift using reference samples. Raw abundance of peptides was normalized by using heavy isotope-labeled internal standards with known peptide concentrations to determine peptide concentration. Raw abundance of glycopeptides was normalized by comparing it with the abundance of 71 non-modified glycopeptides, representing each of 71 proteins from which the glycopeptides monitored were derived. Relative abundance of a glycopeptide was calculated as the ratio of the raw abundance of the glycopeptide to the raw abundance of a peptide from the same glycoprotein. Site occupancy was calculated as the ratio of the raw abundance of any given glycopeptide to the sum of raw abundances of all glycopeptides with the same amino acid sequence. Approximate glycopeptide concentration was calculated by multiplying relative abundance or site occupancy of a glycopeptide by the concentration of a peptide from the same protein. Log-transformed concentration-normalized data for 501 glycopeptides, 452 of which were based on site occupancy and 49 on relative abundance, and for 70 peptide biomarkers were ultimately used for the analysis, totaling 571 unique biomarkers.

Fold changes for individual peptides and glycopeptides were calculated on normalized abundances of control vs. EOC samples, and benign tumor vs. EOC samples. False discovery rate (FDR) was calculated using the Benjamini-Hochberg method (27). Principal component analysis (PCA) was performed on log-concentration-normalized abundances of glycopeptides to investigate differences among the three phenotypes studied. Prior to performing PCA, normalized abundances were scaled such that the distributions of all biomarkers were Gaussian with zero mean and unit variance.

To compare any two phenotypes, age-adjusted linear regression was used on a feature-by-feature basis with phenotype serving as the sole binary independent variable. After correcting for multiple comparisons, differences of any biomarker among phenotype groups compared were considered statistically significant if an FDR of less than 0.05 was reached.

For supervised multivariate modeling, 571 concentration, 49 relative abundance, and 464 site occupancy features were log-transformed and split into a training set consisting of 80% of all samples from women with benign tumors and EOC, and a testing set consisting of the remaining 20% and all healthy controls. To perform binary classification and predict probability of EOC, repeated ten-fold cross-validated least absolute shrinkage and selection operator-regularized logistic regression was used with hyperparameters tuned to prevent overfitting and promote balanced sensitivity and specificity metrics. A subset of 976 of the 1084 total features with low coefficients of variation (<20%) in pooled serum replicates were considered in training binary classification models.

For training a model to distinguish between early- and late-stage EOC, data was split into train and test set at a ratio of 80:20. Prior to training a model, features were scaled so that the distribution had a mean value of 0 and a standard deviation of 1. Logistic regression model with lasso regularization was built using 50 glycopeptide abundance features from a subset of 98/49 markers/pairs that only differed by a single fucose. The probability estimates of a sample in the test set predicted to belong to a particular phenotype was obtained from the trained model.

### Analysis of global glycosylation patterns and site-specific glycoproteomic alterations

Fold change (FC) was determined for each glycopeptide marker by comparing its relative abundance in the studied cohort versus the healthy samples. Markers with statistically significant FC (FDR <0.05) were sorted into markers that were either non-fucosylated or had one fucose residue. Additionally, pairs of markers were identified that differed by a single fucose residue, *i*.*e*., two glycopeptides differing only by fucose. The differential abundances of these markers were calculated and plotted using GraphPad Prism (Boston, USA) and python library matplotlib. 95% confidence intervals (CIs) around the median fold changes were calculated and plotted. Statistical significance was inferred by assessing overlapping CIs and confirming by the numeric p-values. The Kolmogorov-Smirnov test function of GraphPad Prism was used to generate a p-value.

### Lectin staining and microscopy analysis of tumor microarrays

Human tumor microarray slides (Ovarian cancer tissue microarray OV602a slide #28 and #29; BC110118 slide #220 and #221; OV808a slide #52 and #53) were obtained from US Biomax (Derwood, AR, USA). Tumor microarray slides were deparaffinized in xylene and rehydrated in graded anhydrous denatured ethanol sequentially from 100%, 95% to 70%. After tris-buffered saline with 0.1% Tween 20 detergent (TBST) wash (3 times, 5 minutes each), samples were incubated in a carbo-free blocking solution (Vector Laboratories, Burlingame, CA, USA) for 30 minutes to block non-specific binding sites. Slides were then incubated with fluorescein-conjugated lectins. Aleuria Aurantia lectin (Vector Laboratories) was used at a concentration of 5 μg/ml in blocking solution with or without 100 mM L-Fucose (Thermo Fisher Scientific Waltham, MA, USA);

Lens Culinaris Agglutinin (Vector Laboratories) was used at a concentration of 20 μg/ml in blocking solution with or without 100 mM α-methyl-mannoside (Vector Laboratories) for 45 minutes. Slides were washed twice with TBST followed by one wash with deionized water. Mounting media containing DAPI (Abcam) was applied and incubated at room temperature for 5 minutes before placing coverslips. The dried slides were observed with an EVOS M500 imaging system (Thermo Scientific) with the appropriate fluorescent filter. The fluorescence intensity was measured and analyzed using imageJ software, then the mean fluorescent intensity was graphed using GraphPad Prism (GraphPad, San Diego, CA, USA).

### Functional pathway analysis

The QIAGEN⍰ Ingenuity Pathway Analysis (IPA) tools were used on peptide and glycopeptide data for functional interrogation of relevant pathways. Glycopeptide features were ranked by their estimated FDR of differential abundance and mapped to the respective proteins. For the quantitative fold difference for each of these proteins between experiment contrasts, the log fold difference of the most significant glycopeptide mapped to a protein was selected. The IPA Core Expression analysis method was used to identify enriched canonical pathways, potential upstream regulators, and associated protein networks. IPA uses the right-tailed Fisher’s exact test to determine the statistical significance of each reported canonical pathway. Protein log-fold-change was used to calculate the directionality in the IPA analysis. Only genes included in the IPA Knowledge base were used as a reference set in calculating p-values of significance. IPA upstream analysis was used to identify upstream molecule’s regulatory effects that may account for differential expression changes seen in our data, including both transcription and other factors known to affect protein expression. For network inferences, both direct and indirect relationships were considered. Results were filtered by considering relationships described in *Homo sapiens*.

### Cytokine and chemokine quantification

A subset of 65 serum samples (for which sufficient quantities were available) representative of the original set and including 10 benign, 8 stage I, 5 stage II, 34 stage III, and 8 stage IV tumors were randomly chosen for cytokine and chemokine analysis. Analysis was carried out by Rules-Based Medicine (Austin, TX). Samples were thawed at room temperature, vortexed, spun at 3700 x g for 5 min for clarification and transferred to a master microtiter plate. Using automated pipetting, an aliquot of each sample was added to individual microsphere multiplexes of the selected Multi Analyte Profile (MAP) and blocker. This mixture was thoroughly mixed and incubated at room temperature for 1 hour. Multiplexed cocktails of biotinylated reporter antibodies were added robotically and after thorough mixing, incubated for an additional hour at room temperature. Multiplexes were labeled using an excess of streptavidin-phycoerythrin solution, thoroughly mixed and incubated for 1 hour at room temperature. The volume of each multiplexed reaction was reduced by vacuum filtration and washed 3 times. After the final wash, the volume was increased by addition of a buffer for analysis using a Luminex instrument and the resulting data interpreted using proprietary software developed by Rules-Based Medicine. For each multiplex set of analytes, both calibrators and controls were included on each microtiter plate. Eight-point calibrators to form a standard curve were run in the first and last column of each plate and controls at 3 concentration levels were run in duplicate. Standard curve, control, and sample QC were performed to ensure proper assay performance. Study sample values for each of the analytes were determined using 4 and 5 parameter logistics, weighted and non-weighted curve fitting algorithms included in the data analysis package. Results were analyzed on GraphPad Prism by converting resulting levels into *z* scores and plotting these as a heatmap. All values below the lower limit of quantification (LLOQ) were assumed to be half of the LLOQ.

### Gene expression analysis and Cellular Network Knowledge Base analysis

Gene expression data for *MGAT3* and *MGAT4A* for “tumor”; representing malignant EOC, and “metastasis”; representing metastatic tumor deposits, were downloaded from tnmplot.com. Data was plotted and statistical tests was run using GraphPad Prism. Cellular Network Knowledge Base analysis was performed on TCGA Ovarian Cancer data utilizing the CTD^2^ database (https://ctd2-dashboard.nci.nih.gov/dashboard/). Separate queries were run to look for putative interactions (based on mRNA expression data) between glycosyltransferase genes and cytokines/their receptors.

## Results

### Unique glycoproteomic signatures distinguish EOC from benign tumors and fucosylated glycopeptides are associated with EOC

A total of 428 glycopeptides and peptides displayed statistically significant differences in abundance (FDR<0.05) when comparing samples of patients with benign tumors and those with EOC. We performed principal component analysis to assess the segregation between the three phenotypes across first and second principal components. Modest separation between the phenotypes, especially those with EOC, suggested that differential glycoproteomic profiles could be exploited for multivariate model development (figure 1B).

**Figure 1:**
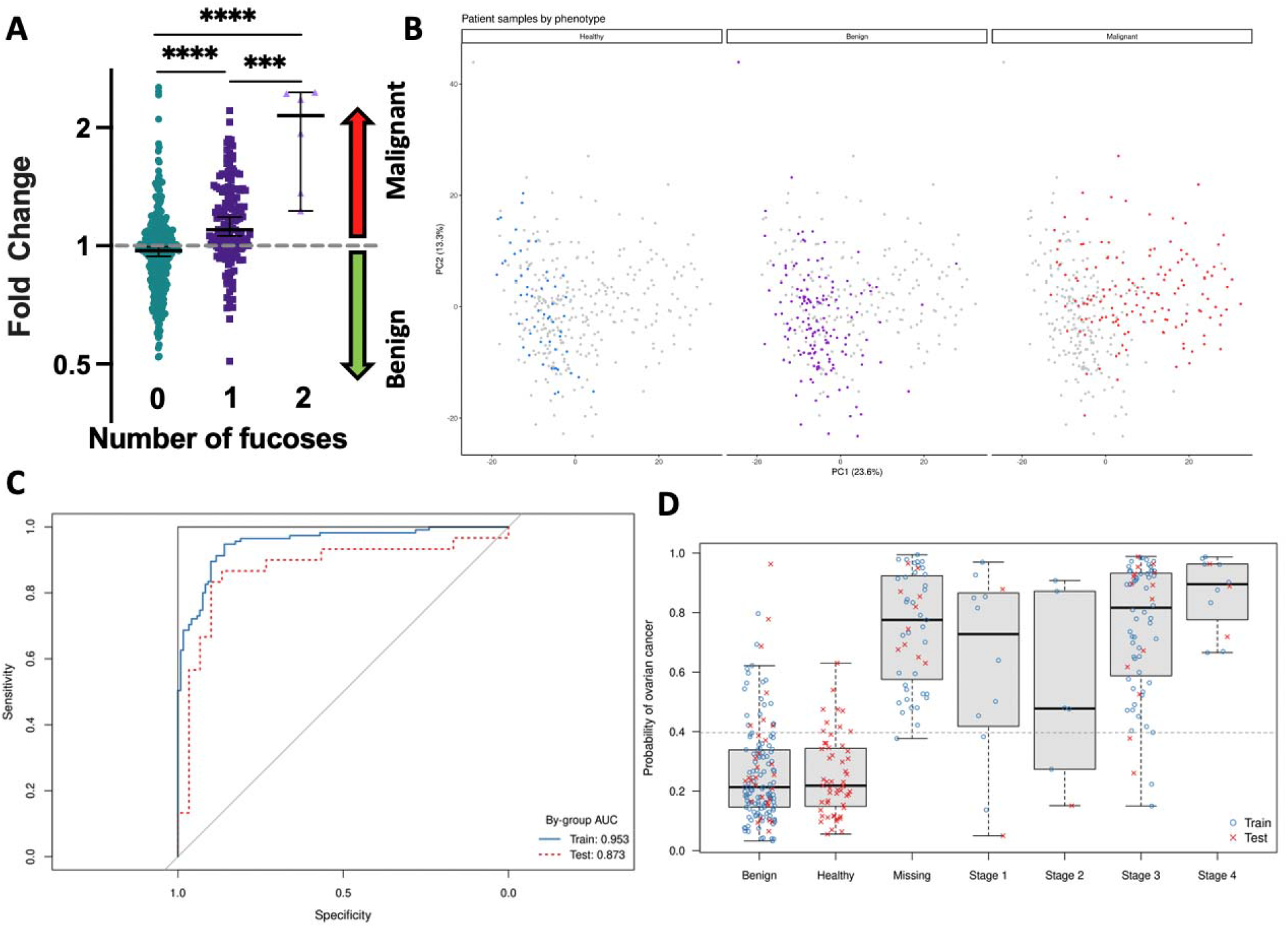
Identification of glycopeptide biomarkers that separate benign and malignant pelvic masses. A) Circulating mono- and di-fucosylated glycopeptide markers are associated with malignant EOC. FC of glycopeptide markers were plotted based on the number of fucose molecules. Solid bars represent the median FC and error bars represent the 95% CI. Non-fucosylated and fucosylated groups have significantly different distributions, and median FCs for mono- and di-fucosylated markers were greater than 1, suggesting an association between fucosylated glycopeptide markers and malignant EOC. Statistical significance between groups was assessed by the Mann-Whitney test. ****indicates p<0.0001 and ***indicates p-value=0.0003. B) Top two principal components in PCA of all 351 subjects included in the analysis with healt y subjects colored blue in the leftmost panel, benign colored purple in the middle panel, and malignant cases colored red in the rightmost panel. C) ROC analysis of multivariate model distinguishing EOC from benign tumors. D) Multivariate model performance by predicted probability, stratified by phenotype and stage.

First, we built a multivariable model to identify EOC (including those with undocumented stage) and benign disease. Five-fold repeated cross-validation in the training set established the optimal LASSO hyperparameter (lambda = 0.045, cross-validated F1 = 0.849). Applying this amount of shrinkage to the panel of 976 features resulted in a logistic regression model that retained 27 biomarkers with non-zero coefficients. The model achieved high accuracy in both the training (accuracy = 0.869, sensitivity = 0.835, specificity = 0.901) and test sets (accuracy = 0.867, sensitivity = 0.867, specificity = 0.867). ROC analysis demonstrated strong performance across a range of cutoffs and little overfitting (training AUC = 0.953; test AUC = 0.873) (Figure 1C). The predicted probability of malignancy increased with cancer stage, and probability distributions were similar across training and test sets (Figure 1D). Notably, when applied to healthy patients not utilized in training, the model resulted in few misclassifications and a spread nearly equivalent to that of the benign tumor cases. The distribution of predicted probabilities indicates a well-trained model, and application to blinded healthy patients and increasing severity with EOC disease progression indicate a link between the glycoproteome and the underlying biology of the disease. As fucosylation has been implicated in a multitude of processes in tumors (28), and fucosylated biomarkers have been reported to be upregulated in the circulation of women with EOC (29,30), we compared glycopeptide fold change (FC) of malignant EOC cases versus benign tumors (figure 1A). Both mono- and di-fucosylated markers had median FCs above 1, suggesting a correlation of these markers with malignant EOC.

### Tri- and tetra-antennary glycans containing fucose are associated with late-stage EOC

Based on the finding that circulating fucosylated glycopeptides were associated with EOC, we asked if there were specific sub-groups of markers or EOC that drove this association. Peptid s carrying tri- and tetra-antennary N-glycans with fucose correlated strongest with late-stage EOC. We then compared the relative abundance of distinct fucosylated glycopeptides in benign tumors, early-stage EOC and late-stage EOC and detected a consistent increase in association with the progression of disease (figure 2C).

**Figure 2:**
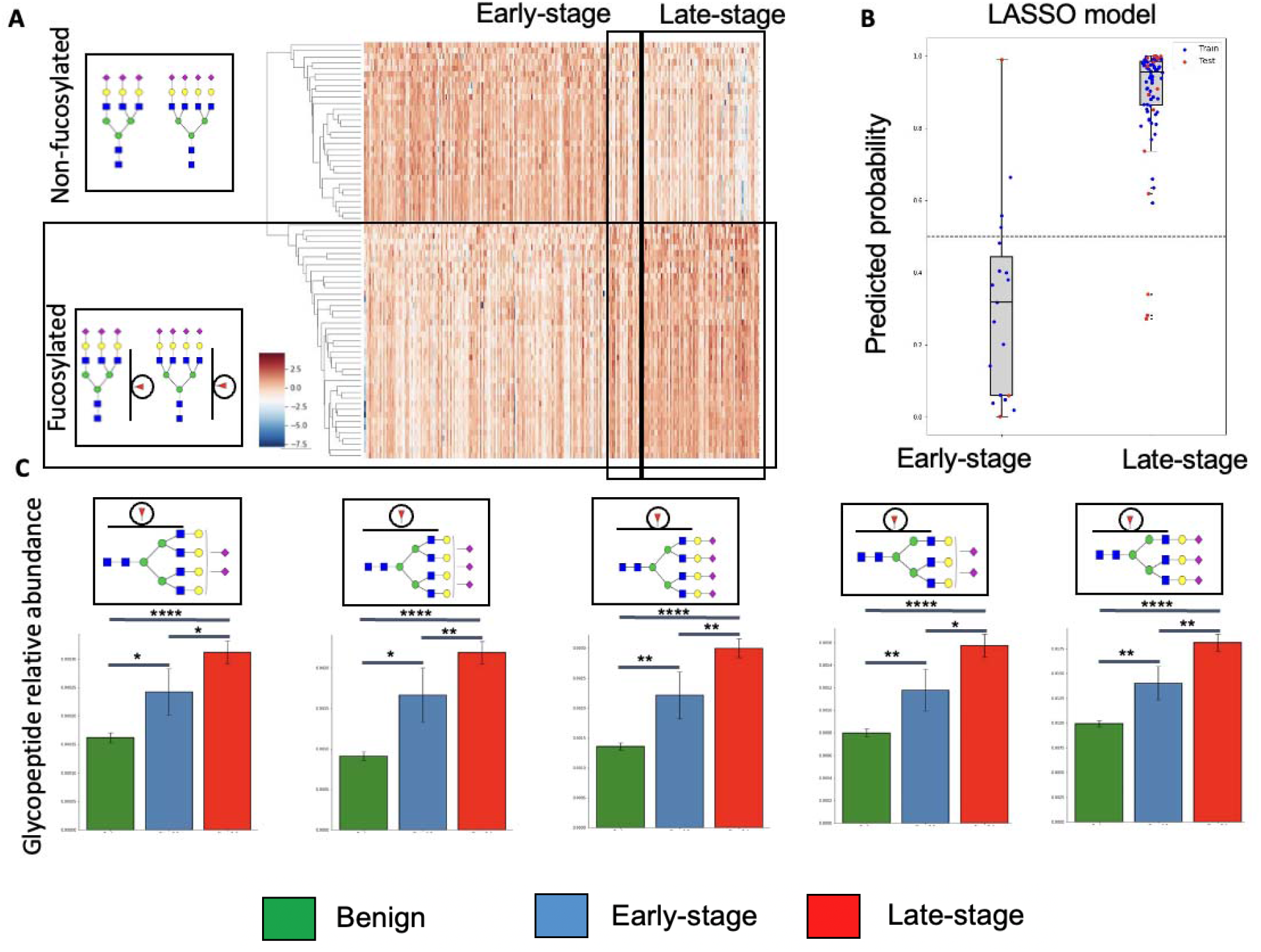
Circulating tri- and tetra-antennary N-glycopeptide markers containing fucose are associated with late-stage EOC. A) Relative abundance of pairs of fucosylated/non-fucosylated markers of all patients separated by pathology are represented as a heatmap. Columns represent patients and rows represent glycopeptide biomarkers. Patients with benign tumors are unmarked, early- and late-stage EOC patients are enclosed in marked black rectangles. Corresponding glycans are indicated with colored geometric SNFG symbols. B) Predicted probability of early- and late-stage classification by a LASSO model built on the pairs of glycopeptide markers represented in figure 2A. C) Bar graph representation of glycopeptide abundance. The three leftmost bar graphs represent glycopeptides with tetra-antennary glycans with varying sialylation. The last two bar graphs represent glycopeptides with tri-antennary glycans with two or three sialic acids. (*p-value<=0.05, **p-value<=0.01, ***p-value<=0.001, ****p-value<=0.0001)

To further this finding, we isolated pairs of tri- and tetra-antennary N-glycopeptides that only differed by a single fucose and calculated their relative abundances (represented as a heatmap in figure 2A and defined in supplementary information). The fucosylated form of these markers was associated with late-stage EOC. To assess if these pairs of markers differentiated between early- and late-stage EOC, we developed a logistic regression model with LASSO regularization. The model achieved high accuracy in both the training (accuracy = 0.963, sensitivity = 1, specificity = 0.812) and test sets (accuracy = 0.8, sensitivity = 0.824, specificity = 0.667) and identified early and late-stage disease (figure 2B).

### Metastasis from EOC displays higher levels of surface fucosylation and gene expression patters suggestive of tri- and tetra-antennary glycans

Given the fucosylation differences observed in circulating glycoproteins, we assessed fucosylation levels by staining tumor tissue microarrays with AAL, a fucose-binding lectin. Metastatic EOC tissue derived from metastasis to the omentum and peritoneum had significantly higher levels of AAL staining (figure 3A-B). We confirmed these findings by using another fucose-binding lectin LCA (supplementary figure 1 and 2).

**Figure 3:**
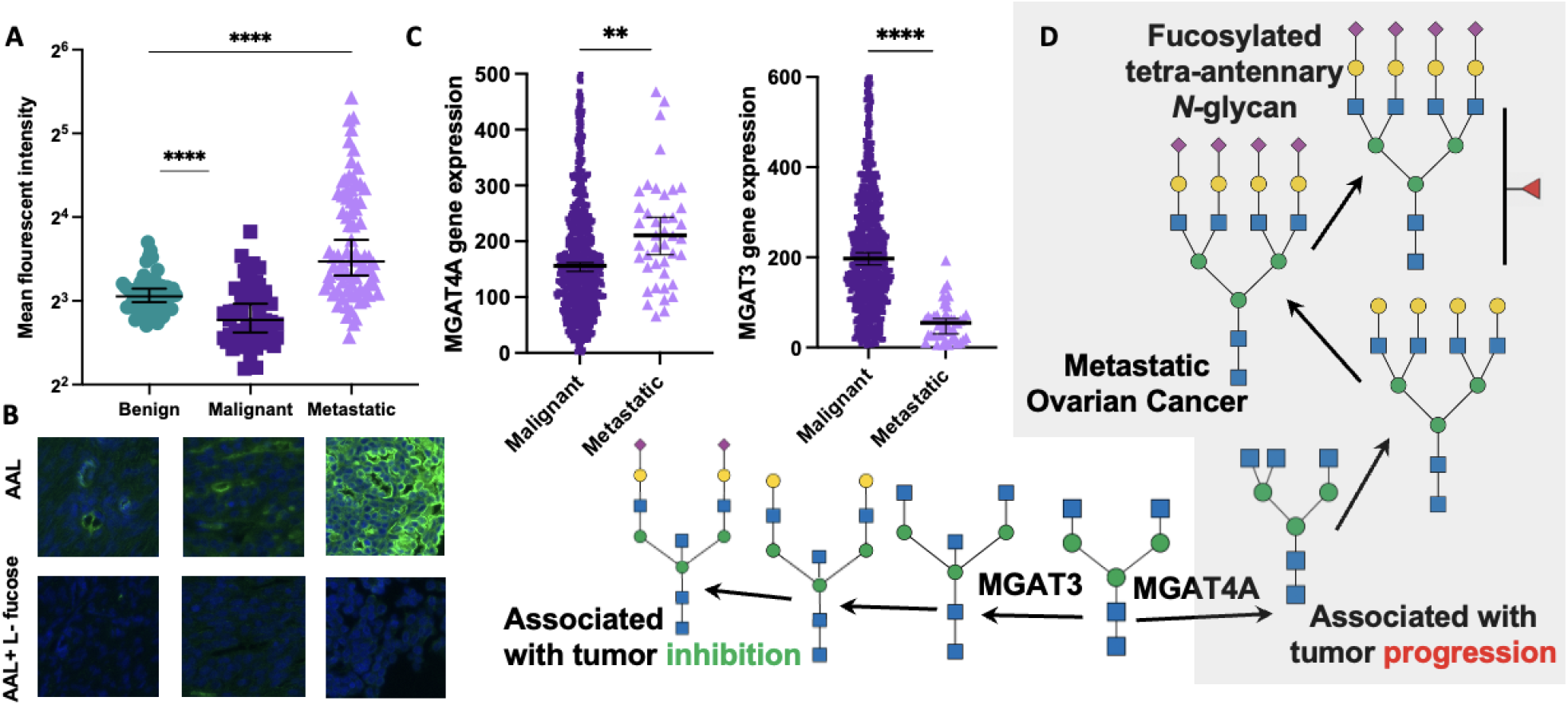
Glycosylation profiles in malignant and metastatic EOC tissues. A) Quantification of staining with AAL. Individual points represent mean fluorescence intensity of individual tissue on a tumor microarray. Solid bars represent the median of the histological group, and error bars represent the 95% CI. p-value was calculated by Kolmogorov-Smirnov test. ****indicates p<0.0001. B) Representative images of AAL staining. The upper panels show staining with AAL. In the lower panels, tissues were incubated with the lectin AAL and L-fucose, as a control. C) Metastatic tissue shows higher levels of *MGAT4A* and lower levels of *MGAT3* mRNA expression. Mann-Whitney’s test used to assess statistical significance. **represents a p-value of 0.0013 and ****represents a p-value <0.0001 D) Pathway of N-glycan extension showing the association of MGAT4A upregulation and MGAT3 downregulation with tumor progression (31).

Circulatory changes of increased branching leading to higher levels of tri- and tetra-antennary glycans we describe in the previous section have also been described to occur in the tumor glycocalyx of advanced tumors (31). Many of these changes are driven by changing levels of the glycosyltransferases MGAT3 and MGAT4A. We therefore analyzed gene expression levels of glycosyltransferases involved in the formation of tri- and tetra-antennary glycans in metastatic tissue and malignant tumors. Metastatic tissue exhibited higher expression of *MGAT4A*, a glycosyltransferase involved in forming tri-antennary branches by adding a β1-4 N-acetylglucosamine (GlcNAc), and lower expression levels of *MGAT3*, an enzyme involved in adding a bisecting GlcNAc that may reduce terminal modifications of N-glycans (figure 3C-D)(32). Similar expression patterns occurred in the primary tumor as well in advanced stage EOC (supplementary figure 3).

### Circulating cytokines are associated with tri- and tetra-antennary N-glycans containing fucose

Given that similar glycosylation patterns were detected in blood and in tissues of late-stage EOC, we posited that common factors might induce overlapping glycosylation programs in hepatocytes, which are the main source of markers included in the glycoproteome panel, and in the tumor cells. We quantified a panel of cytokines and chemokines in early and late-stage EOC samples (figure 4A). Of these, IL-6, IL-8, IL-10 and MCP-1 exhibited a general upregulation in late-stage EOC. Notably, CA125, CA19-9 and CEA were slightly higher in late-stage EOC, but did not seem to consistently indicate progression of disease.

**Figure 4:**
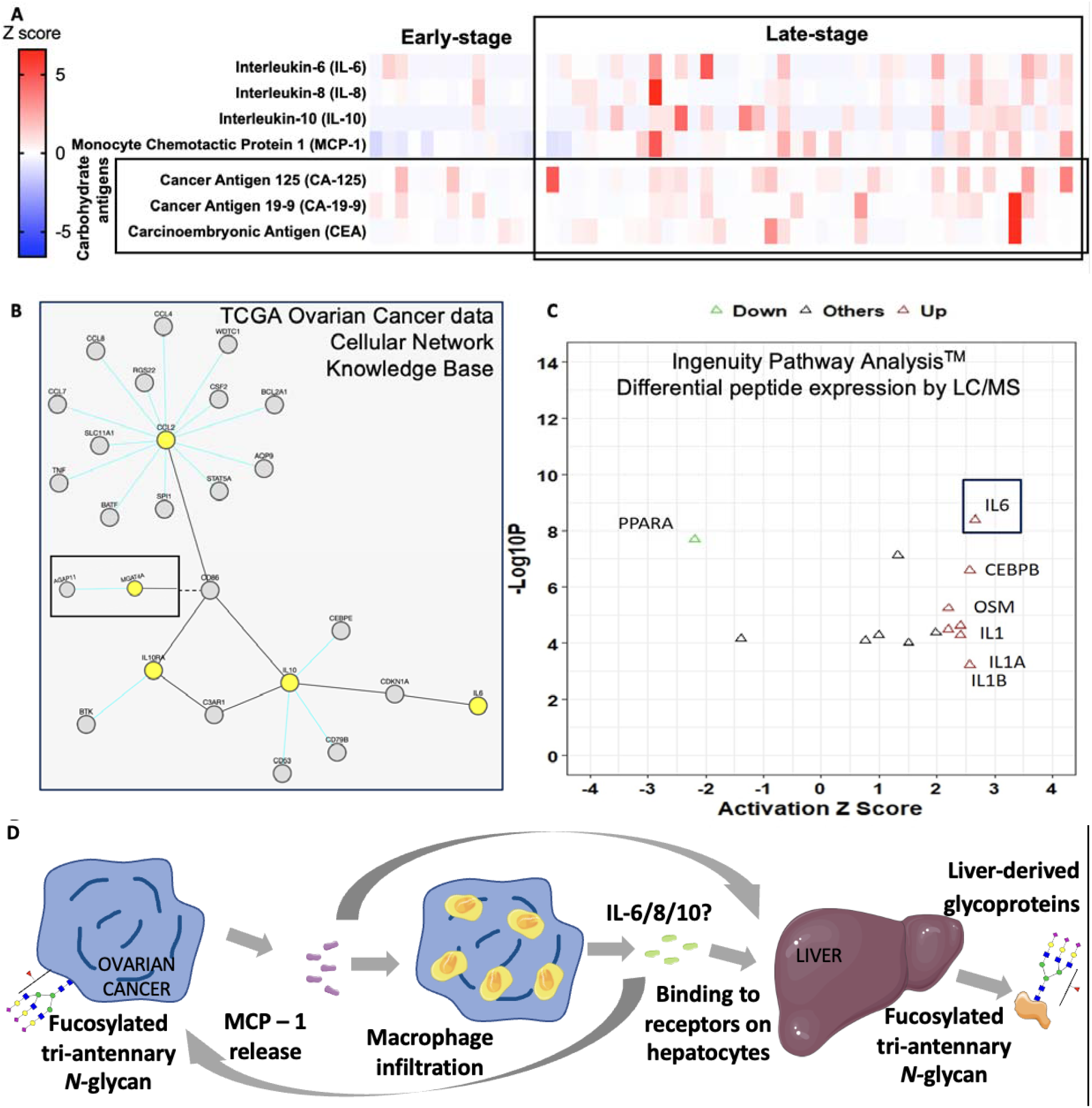
Possible drivers of common glycosylation programs in hepatic-derived glycoproteins and tumor glycoproteins. A) Heatmap representation of z-scores cytokines and cytokines showing generally higher levels in late-stage EOC. B) Cellular Network Knowledge Base analysis. Analysis performed on the TCGA Ovarian Cancer database shows a putative network leading from MCP-1 (CCL2) to MGAT4A. Final analysis included here shows the composite of two analyses: one with IL6, IL10, and CCL2, and the other with these genes along with *MGAT3* or *MGAT4A*. Of the last two genes mentioned, only *MGAT4A* had interactions with the cytokine network. For convenience, the interactions of IL6, IL10, CCL2, and *MGAT4A* are graphically represented as a composite image. C) Ingenuity Pathway Analysis on peptide-level data points to IL-6 as a potential upstream regulator of changes detected in this study. Y-axis represents level of statistical significance and X-axis represents activation z score. D) Hypothetical pathway leading to comparable alterations in protein glycosylation at the tumor site and in liver-derived glycoproteins.

To understand if these factors were related to the gene expression profiles, we queried the Cellular Network Knowledge Base for potential interaction between the cytokines, *MGAT3* and *MGAT4A*. These glycosyltransferases were chosen given the previous observation that they were significantly different in metastatic EOC tissue. Strikingly, both IL-6 and IL-10 were related to *MGAT4A* expression via CD86 and CCL2 (figure 4B).

We then searched for upstream regulators using functional pathway analysis based on differential peptide expression, and identified IL-6 as a primary hit (figure 4C and supplementary table 1).

## Discussion

In this study, we analyzed peripheral blood samples using a novel technology platform consisting of LC-MS coupled with a high throughput data processing engine (33) that allows scalable characterization of the glycoproteome of clinical samples. We identified glycopeptide markers expressed at higher levels in blood of EOC patients compared to patients with benign tumors and healthy controls. We generated a classifier that allowed the identification of patients presenting with malignant masses with high specificity and sensitivity. We used a similar approach to build a model that differentiated between early- and late-stage EOC. As early triaging and noninvasive staging influence treatment protocols for women with pelvic masses, these data that demonstrate reliable differentiation of benign versus malignant tumors with ability to detect early stage disease are an important advancement in EOC diagnostics.

The biomarkers described herein represent glycosylated variants of relatively abundant blood proteins and may be the product of a systemic response to the disease. Due to intrinsic amplification effects, possibly mediated by cytokines and chemokines, our analytical platform allows for early stage diagnosis. This differs categorically from tests such as CA-125, CA19-9, cell-free DNA and circulating tumor DNA (34), that detect molecules produced at the tumor site and rely on their accumulation in blood for detection of disease.

Fucosylation appeared to be a key feature of biomarkers in malignant EOC. As an advancement of earlier glycomic (35,36) nd glycoproteomic studies of EOC (37–41), we detected high levels of fucosylation combined with tri- and tetra-antennary glycans in late-stage EOC. Similar findings have been described in earlier glycomic studies of serum glycoproteins (42) and in ascitic fluid as well (36). As protein fucosylation has been described to modulate important mechanisms including immune cells recognition and cell-cell interactions (43), we speculate that the changes observed in our glycoprotein panel may be implicated in disease progression.

Moreover, our observation that overlapping glycosylation changes occur in liver-derived glycoproteins and in late-stage EOC tissues suggest that they may be driven by common mechanisms. We hypothesize that factors including MCP-1, IL-6, IL-8, and IL-10 are released in the tumor microenvironment during disease progression and promote cycles of infiltration of myeloid cells and increasing inflammation. This dynamic is accompanied by the modulation of expression of glycosyltransferases, including *MGAT3* (44–47) and *MGAT4A*, at the tumor site and in the liver that may lead to increased decoration of both circulating glycoproteins as well as tumor-surface proteins with tri- and tetra-antennary glycans containing fucose.

Limitations of our study include: 1) its retrospective design, thus requiring further prospective validation. We have initiated a study (NCT03837327: Clinical Validation of the InterVenn Ovarian CAncer Liquid Biopsy, VOCAL) to investigate the diagnostic utility of these circulatory glycoproteomic markers in the triage of newly diagnosed adnexal masses. 2) the limited number of early-stage EOC samples that affect modeling that differentiates between early- and late-stage EOC. This drawback also would be mitigated by a large prospective cohort. Additionally, it would be important to study matched tumor and blood samples using the same analytical platform to confirm the correlation in glycosylation profiles in tumors and in circulating glycoproteins.

In conclusion, our study demonstrates the relevance and feasibility of mass spectrometric analysis of liquid biopsies not only as a research tool for the identification of novel biomarkers, but also for their application as clinically actionable diagnostic tests. Additionally, we provide initial evidence related to the mechanisms that induce the observed glycosylation signatures. Future studies may also shed light on the potential roles of the glycosylation signatures in disease progression.

## Data Availability

All data produced in the present work are contained in the manuscript

## Acknowledgments

The authors thank Ranjan Bhadra for sample management and Cassie Xu for her contributions in establishing protocols to analyze mRNA sequencing data. We thank Dr. Carolyn Bertozzi and Dr. Nissi Varki for their suggestions regarding study methodologies. We are also grateful to Dr. Carlito Lebrilla for reviewing the manuscript and providing feedback.

## Author contributions

The project was conceptualized by CD, DS, KL, and FS. Data was generated and analyzed by CD, PR, GX, CP, RR, BZ, AS, PA, CC, FJ. Data was visualized by CD, PR, CP, BZ, PA, and FJ. KM acquired clinical samples. The manuscript was written by CD and KL and edited by all authors.

## Definition of tri- and tetra-antennary N-glycans

In this study, we call any glycan with three antennas/branches (as shown in the figure below) as a tri-antennary glycan. Similarly, if there are four antennas/branches, it is tetra-antennary. Independent of the number of antennas, an additional fucose (red triangle) can be bound to represent tri-antennary or tetra-antennary N-glycans containing a fucose.

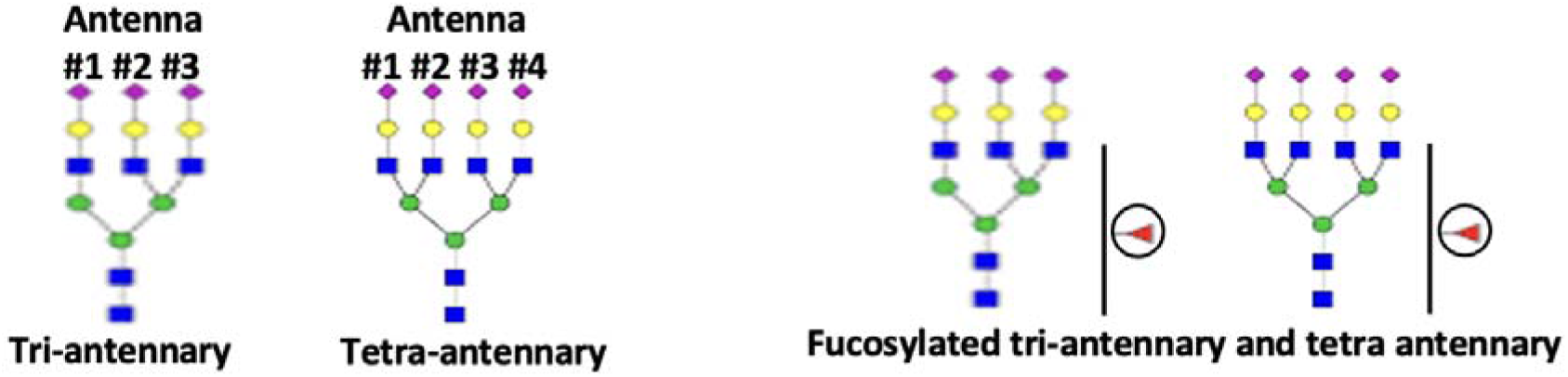

**Supplementary figure 1:**
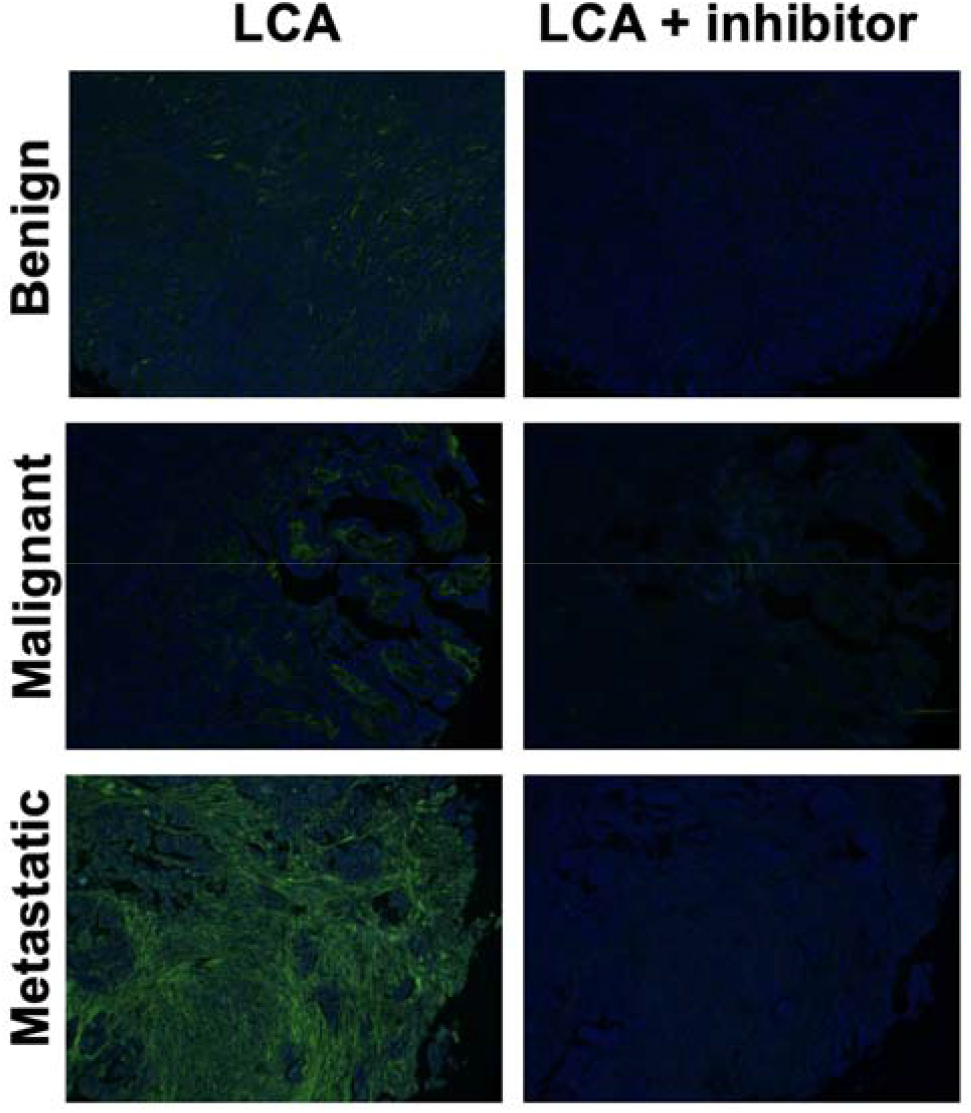
Representative images of LCA staining on Ovarian cancer tumo microarrays.

**Supplementary figure 2:**
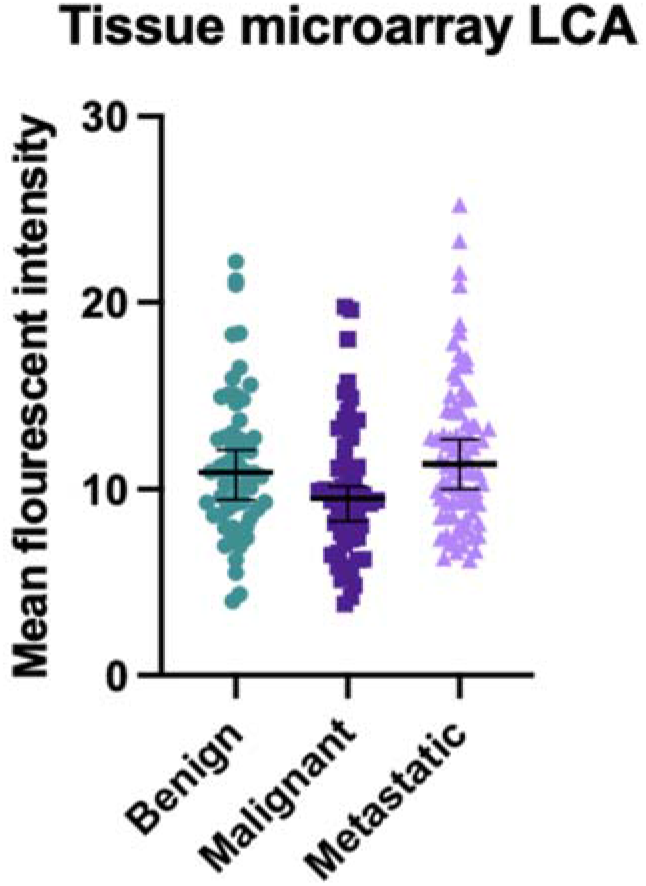
Quantification of LCA fluorescence on Ovarian cancer tumor microarrays.

**Supplementary figure 3:**
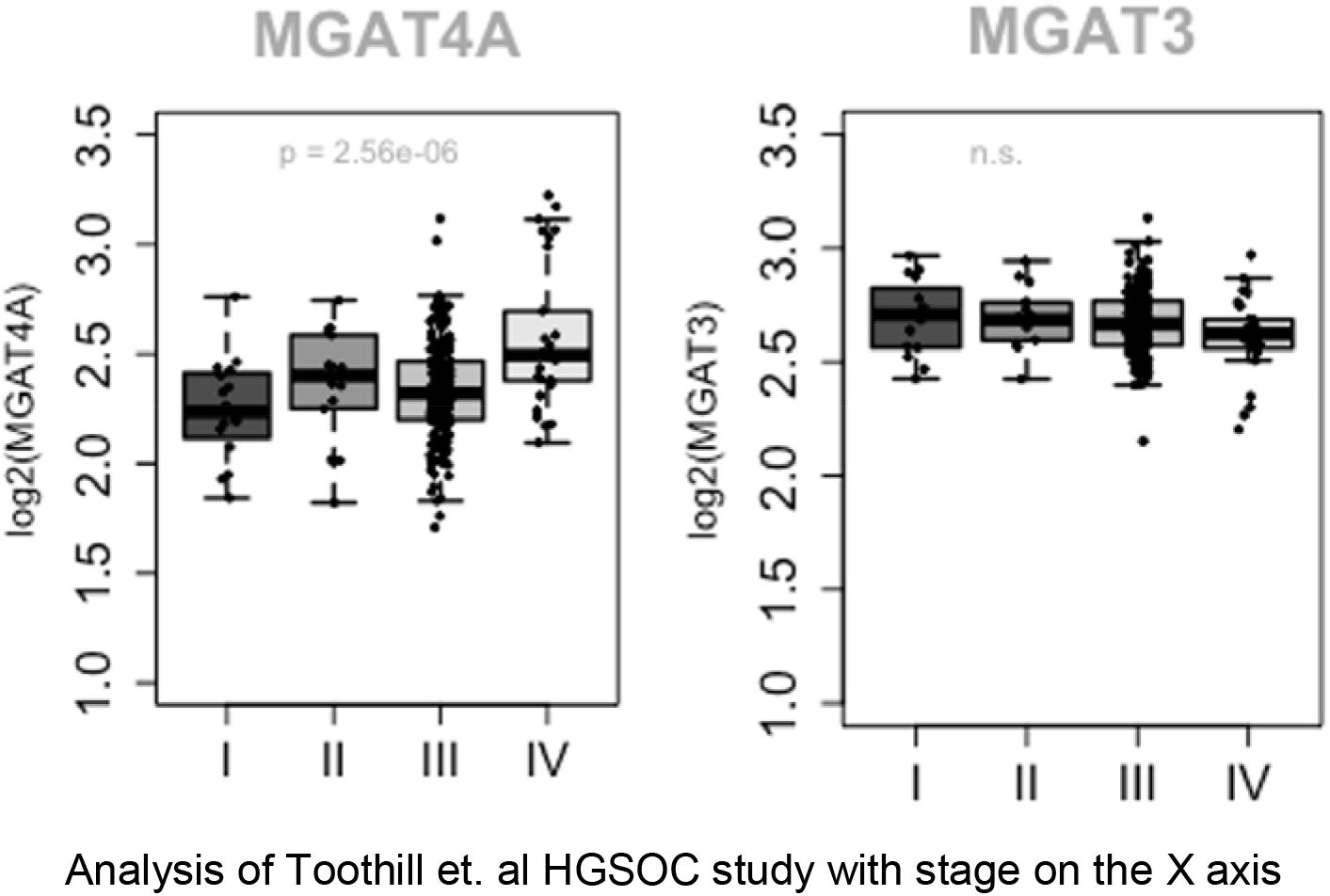
mRNA gene expression of MGAT4A and MGAT3 by stage of EOC.

**Supplementary Table 1:** Table of Top 20 Predicted Upstream Regulators by QiAGEN Ingenuity Pathway Analysis (IPA) Upstream regulator prediction tool. The table includes the mean pValue of prediction, the endogenous molecule type and the target molecules observed in our study data across comparisons. The mean pValue is the average of p-values of predictions of the upstream regulator in the benign disease vs healthy, early disease vs healthy and the late disease vs healthy study comparisons.

| Regulator | pValue | Type | Targets |
| --- | --- | --- | --- |
| HNF1A | 8.60E-11 | transcription regulator | AHSG,APOH,APOM,C1S,C4BPA,ITIH4,SERPINA1,SERPIN G1,VTN |
| IL6 | 8.80E-08 | cytokine | AGT,APOB,CLU,HP,ORM1,SE RPINA1,SERPINA3 |
| HNF4A | 1.28E-06 | transcription regulator | AGT,APOB,APOC3,APOM,SE RPINA1 |
| SREBF1 | 8.29E-06 | transcription regulator | CFI,MT-CO2,SERPINA1,SERPINA3,T F |
| PPARA | 0.000226 | ligand-dependent nuclear<br>receptor | APOC3,APOM,MT-CO2 |
| RXRA | 0.000238 | ligand-dependent nuclear<br>receptor | APOC3,APOM,KNG1 |
| NR1H3 | 0.000273 | ligand-dependent nuclear<br>receptor | APOC3,APOM |
| IL22 | 0.000559 | cytokine | HP,SERPINA3 |
| TCF | 0.00057 | group | APOD,SERPINA1,SERPINA3 |
| SMARCA4 | 0.000824667 | transcription regulator | A2M,AGT,AZGP1,CP,FN1,IGH<br>G1 |
| CTNNB1 | 0.000924667 | transcription regulator | APOD,FN1,SERPINA1,SERPI<br>NA3 |
| GFI1 | 0.000941333 | transcription regulator | SERPINA1,SERPINA3 |
| CEBPB | 0.00114 | transcription regulator | APOB,FN1,HP |
| NR5A2 | 0.001756667 | ligand-dependent nuclear<br>receptor | APOM,HP |
| COL5A1 | 0.002063333 | other | FN1 |
| GBP2 | 0.002063333 | enzyme | FN1 |
| ITGB5 | 0.002063333 | other | VTN |
| NUP153 | 0.002063333 | other | THRB |
| PLPP3 | 0.002063333 | phosphatase | FN1 |
| PPP2R5E | 0.002063333 | phosphatase | FN1 |

